# Persistent *IDH* mutations are not associated with increased relapse or death in patients with *IDH*-mutated acute myeloid leukemia undergoing allogeneic hematopoietic cell transplant with post-transplant cyclophosphamide

**DOI:** 10.1101/2023.08.14.23294087

**Authors:** Niveditha Ravindra, Laura W. Dillon, Gege Gui, Matthew Smith, Lukasz P. Gondek, Richard J. Jones, Adam Corner, Christopher S. Hourigan, Alexander J. Ambinder

**Affiliations:** Laboratory of Myeloid Malignancies, Hematology Branch, National Heart Lung, and Blood Institute, National Institutes of Health, Bethesda, MD; Sidney Kimmel Comprehensive Cancer Center, Johns Hopkins University, Baltimore, MD; Bio-Rad Laboratories, Digital Biology Group, 5731, W. Las Positias Blvd, Pleasanton, CA; Myeloid Malignancies Program, National Institutes of Health, Bethesda, MD

## Abstract

The presence of measurable residual disease (MRD) prior to an allogeneic hematopoietic transplant (alloHCT) in Acute Myeloid Leukemia (AML) has been shown to be associated with an increased risk of post-transplant relapse. Since the Isocitrate Dehydrogenase genes (*IDH1*/*2*) are mutated in a considerable proportion of patients with AML, we studied if these mutations would serve as useful targets for MRD. Fifty-five *IDH*-mutated AML patients undergoing non-myeloablative alloHCT with post-transplant cyclophosphamide at a single center were sequenced at baseline using a multi-gene panel followed by targeted testing for persistent *IDH* mutations at the pre- and post-alloHCT timepoints by digital droplet PCR or error-corrected next generation sequencing. The cohort included patients who had been treated with *IDH* inhibitors pre- and post-transplant (20% and 17% for *IDH1* and 38% and 28% for *IDH2*). Overall, 55% of patients analyzed had detectable *IDH* mutations during complete remission prior to alloHCT. However, there were no statistically significant differences in overall survival (OS), relapse-free survival (RFS), and cumulative incidence of relapse (CIR) at 3 years between patients who tested positive or negative for a persistent *IDH* mutation during remission (OS: *IDH1* p=1, *IDH2* p=0.87; RFS: *IDH1* p=0.71, *IDH2* p= 0.78; CIR: *IDH1* p=0.92, *IDH2* p=0.97). There was also no difference in the prevalence of persistent *IDH* mutation between patients who did and did not receive an *IDH* inhibitor (p=0.59). Mutational profiling of available relapse samples showed that 8 out of 9 patients still exhibited the original *IDH* mutation, indicating that the *IDH* mutations remained stable through the course of the disease. This study demonstrates that persistent *IDH* mutations during remission is not associated with inferior clinical outcomes after alloHCT in patients with AML.

---

In patients with acute myeloid leukemia (AML) there is considerable evidence that suggests that measurable residual disease (MRD) detection during and after treatment (1), and particularly before allogeneic hematopoietic cell transplantation (alloHCT), predicts subsequent relapse and inferior survival outcomes (2, 3). ELN guidelines recommend the use of validated molecular tests for AML MRD where available, but the appropriate targets for such monitoring are incompletely defined (4). Mutations in the epigenetic regulator Isocitrate Dehydrogenase (*IDH*) genes are observed in approximately 20% of patients diagnosed with AML (5). AlloHCT may improve outcomes for such patients (6, 7). To further investigate the utility of persistent *IDH1* and *IDH2* mutations as AML MRD markers, we screened for the detection of these variants at pre- and post-transplant remission timepoints in a cohort of AML patients undergoing alloHCT.

Consecutive adult patients between 2015 and 2020 with *IDH*-mutated AML who achieved complete remission (CR) and underwent non-myeloablative alloHCT with cyclophosphamide-based graft versus host disease prophylaxis at the Johns Hopkins Sidney Kimmel Comprehensive Cancer Center were included in this study. Patient characteristics and outcomes for this cohort have been previously reported (8) although 5 patients were excluded because samples were not available **(Supplementary Figure 1)**. Written and informed consent was obtained from all patients and the study was approved by the internal ethics committee in accordance with the Declaration of Helsinki. Baseline and relapse BM DNA samples were sequenced using the 75-gene VariantPlex myeloid panel (ArcherDx, Boulder, CO). Assessment for persistent *IDH* mutations was performed on pre- and post-alloHCT remission samples using droplet digital PCR (ddPCR) or a custom error-corrected targeted next generation sequencing panel (ecNGS). Additional clinical and testing details are provided in **Supplementary Data 1-5**.

Baseline DNA samples for sequencing were available for 55 patients. Clinical characteristics are described in **Supplementary Table 1**. An exclusive *IDH1* or *IDH2* mutation was identified in 26 patients each, while the remaining 3 patients had a mutation detected in both genes. A minority of patients in the *IDH1* (20% and 17%) and *IDH2* cohorts (38% and 28%) received an *IDH* inhibitor pre- or post-alloHCT. Only two patients experienced non-relapse mortality after transplantation.

In the *IDH1*-mutated cohort, the median variant allele frequency (VAF) at diagnosis was 35% (range: 3%-49%). We identified R132H and R132C in 44.8% of samples each followed by R132G in 10%, and R132L in 3.4% of samples. One sample harbored double mutations with R132L and R132C. *NPM1, DNMT3A, FLT3, SRSF2* and *PTPN11* were most commonly co-mutated with *IDH1* **(Supplementary Figure 2)**. In the *IDH2*-mutated cohort, the R140Q mutation was most common (72.4%), followed by R172K (20.6%) and R140W (7%). The median VAF at diagnosis for the R140 and R172 hotspots were 38% and 31%, respectively (p=0.65) (range: 0.4%-49% and 0.4%-47%). *DNMT3A, FLT3, SRSF2, NPM1*, and *BCOR* were the top co-mutated genes with *IDH2* **(Supplementary Figure 2)**.

It has previously been reported that the persistence of *IDH1* R132 and *IDH2* R172 mutations in remission may be associated with post-transplant relapse (9, 10). We performed targeted assessment by ddPCR and/or ecNGS on 49/55 patients who had sufficient DNA available at the pre-alloHCT timepoint, and 27 samples (55%) tested positive for a persistent *IDH* mutation. The median VAF in the *IDH1* R132 positive samples was 5.3% (range: 0.15%-46%) and for the *IDH2* R140 positive samples was 4.4% (range: 0.19%-48%). There was a single *IDH2* R172 positive patient with a VAF of 0.78%. There was no significant difference in the VAFs in pre-transplant remission samples between *IDH1* R132 and *IDH2* R140 hotspots (p=0.86). Overall, 8/13 patients who received an *IDH* inhibitor before alloHCT and were tested had a detectable persistent *IDH* mutation. There was no significant difference in the prevalence of pre-transplant *IDH* mutation persistence between those who did and did not receive an *IDH* inhibitor (p=0.59). Similar results have been observed previously, with *IDH* mutation clearance at CR achieved in 41% and 30% of patients who received ivosidenib and enasidenib, respectively (11). The overall survival (OS), relapse-free survival (RFS), and cumulative incidence of relapse (CIR) were not significantly different between the patients that showed *IDH* mutation persistence in remission and those that did not, for *IDH1* (OS: 93% vs 81% at 3 years, p=1; RFS: 77% vs 74% at 3 years, p=0.71; Relapse: 16% vs 26% at 3 years, p=0.92) and for *IDH2* (OS: 63% vs 60% at 3 years, p=0.87; RFS: 57% vs 56% at 3 years, p=0.78; Relapse: 29% vs 30% at 3 years, p=0.97) **(Figure 1A-C)**. All 6 patients who had persistent pre-alloHCT *IDH* mutation but received a post-alloHCT *IDH* inhibitor maintained remission, while 6/21 patients with persistent pre-alloHCT *IDH* mutation who did not receive a post-alloHCT inhibitor relapsed (p=0.28).

**Figure 1A-C:**
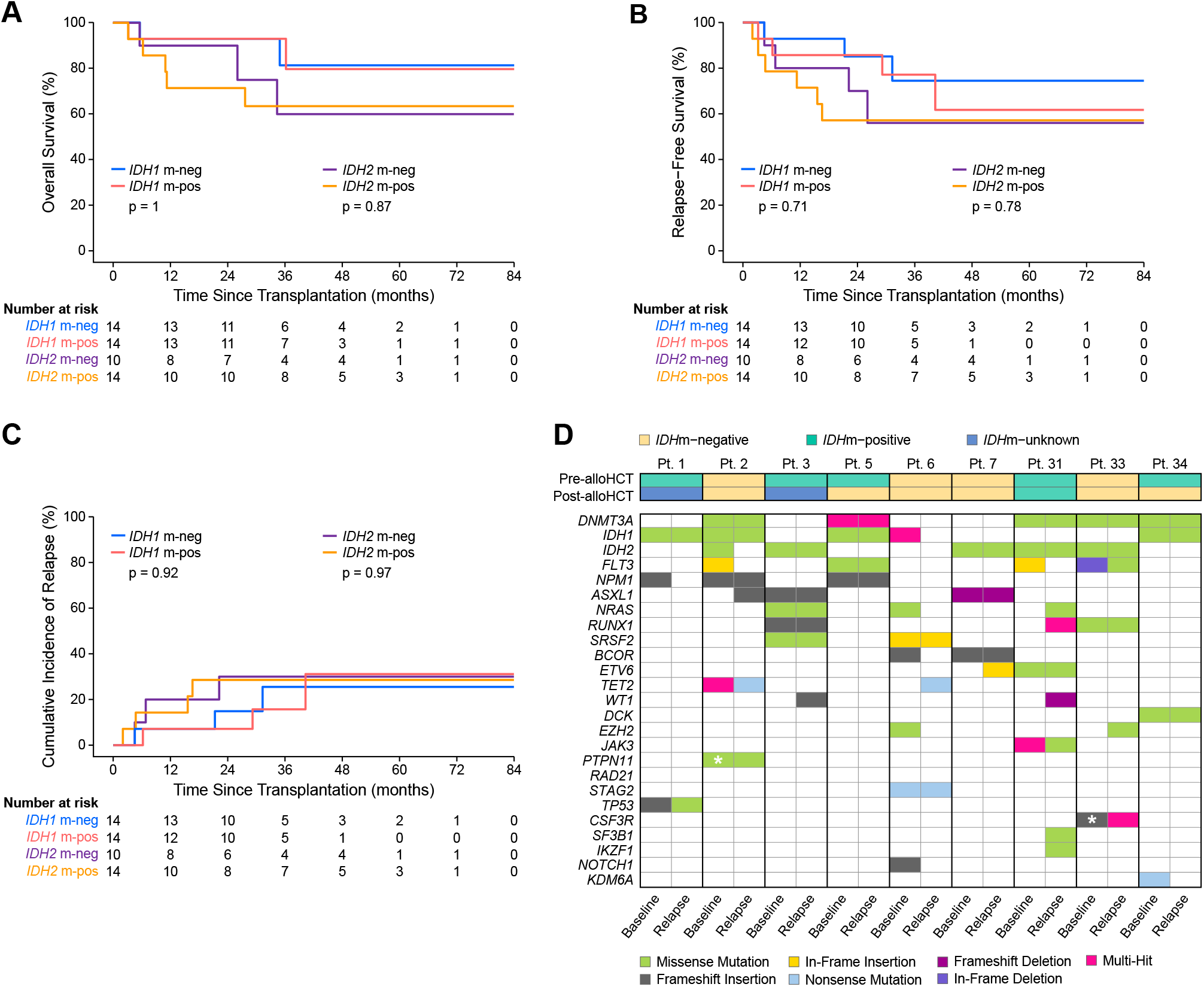
Overall survival (%), Relapse free survival (%) and Cumulative Incidence of Relapse (%) were not significantly different in patients with and without persistent *IDH* mutation for *IDH1* (n=28) or *IDH2* (n=24) tested prior to alloHCT. **Figure 1D:** Comparison of variants identified at baseline and relapse in 9 patients. The ‘*’ indicates mutations present at very low variant allele frequency in baseline samples that were not identified during the initial analysis. alloHCT, allogeneic hematopoietic cell transplant; Pt, patient; m-neg, mutation-negative; m-pos, mutation-positive.

At the post-alloHCT timepoint, 50 patients were assessed by ddPCR/ecNGS. Five patients had persistent *IDH* mutations detectable (10%); 3 had an R140Q and 2 had an R132 mutation. Only two of these patients relapsed and died while the remaining three remained in remission.

In the overall cohort, 12 of 55 patients (22%) relapsed after alloHCT. Only two of five patients positive by clinical flow cytometry MRD testing prior to alloHCT subsequently relapsed (**Supplementary Table 2)**. Two patients who relapsed had received an *IDH* inhibitor before transplantation, one of whom tested positive for persistent *IDH* mutation at the pre-alloHCT timepoint. BM samples from the time of relapse were available for 9 of these 12 patients, sequencing revealed the original *IDH* mutation seen at initial diagnosis in 8 patients indicating that these mutations remain stable through the course of the disease. Prior studies have indicated that these mutations are acquired early during leukemogenesis and are frequently identified in the ancestral AML clone (12). The patient who had an *IDH-*mutation-negative relapse had two *IDH1* mutations at baseline but tested negative for both mutations pre-alloHCT, post-alloHCT, and at relapse. While some of the other mutations identified at baseline (in the *BCOR, EZH2*, and *NRAS* genes) were also absent, the original *SRSF2* and *STAG2* mutations were still present in the relapse sample. In the remaining patients, 19 additional mutations with VAFs ranging from 1.2%-49% were identified. *WT1* (n=3), *PTPN11, NRAS, RUNX1*, and *CSF3R* (n=2 each) were the most common new mutations acquired at relapse. Two of these mutations were present in the baseline sample at very low VAFs but were filtered out during the initial analysis **(Figure 1D)**.

In conclusion, we were unable to demonstrate that *IDH* mutation persistence in remission is associated with an increased risk of relapse or poor survival in adults with AML undergoing non-myeloablative alloHCT with post-transplant cyclophosphamide. This work highlights that detection of an AML-associated somatic mutation in remission is not necessarily representative of leukemic persistence or increased relapse risk. Although patients in our single-center cohort underwent similar pre-alloHCT regimens, a major limitation of this work is the sample size and the small number of relapses in each group. Our cohort also had several patients who had *IDH* inhibitor therapy that was initiated both pre- and post-alloHCT, which may have altered the clinical implications of persistence or clearance of *IDH* mutations. Larger cohorts of patients with *IDH* mutated AML undergoing alloHCT will be required to validate and generalize these findings.

## Data Availability

Raw sequencing data generated during this study has been deposited in the NCBI Sequence Reads Archive (PRJNA998386).

## Acknowledgements

This work was supported by the Intramural Research Program of the National Heart, Lung, and Blood Institute and grants from the National Institutes of Health/National Cancer Institute (P01 CA225618, P30 CA06973). The Center for Cancer Research Genomics core provided support for digital droplet PCR experiments.

## Notes

**Conflict of Interest** The National Heart, Lung, and Blood Institute receives research funding for the laboratory of CSH from the Foundation of the NIH AML MRD Biomarkers Consortium. AC is an employee and shareholder in Bio-Rad Laboratories. All other authors report no relevant conflict of interest.

### Competing Interest Statement

The National Heart, Lung, and Blood Institute receives research funding for the laboratory of CSH from the Foundation of the NIH AML MRD Biomarkers Consortium. AC is an employee and shareholder in Bio-Rad Laboratories. All other authors report no relevant conflict of interest.

### Author Declarations

Approval to conduct this study was obtained from Johns Hopkins Sidney Kimmel Comprehensive Cancer Center: IRB00223526 (Development of a Registry of Patients with Myeloid Malignancies), IRB00357598 (Assessment of Remission Depth and Prognostic Impact of Molecular MRD in Patients with Myeloid Malignancies)

