## Supplementary material for "Persistent *IDH* mutations are not associated with increased relapse or death in patients with *IDH*-mutated acute myeloid leukemia undergoing allogeneic hematopoietic cell transplant with post-transplant cyclophosphamide": Data Supplement

### Supplementary Data

#### 1. Patients and samples

Sixty-one consecutive *IDH*-mutated acute myeloid leukemia (AML) patients who underwent allogeneic hematopoietic cell transplantation (alloHCT) between 2014 and 2020 at the Johns Hopkins Sidney Kimmel Comprehensive Cancer Center were initially included in the study. 5 patients who had unavailable baseline DNA for sequencing and 1 patient who was not in complete remission prior to alloHCT were excluded. Pathogenic *IDH* mutations were defined as mutations resulting in amino acid changes at or adjacent to the *IDH1* R132 residue or the *IDH2* R140 or R172 residues. All patients received the same non-myeloablative conditioning regimen consisting of cyclophosphamide 14.5mg/kg/day intravenously (IV) on days -6 and -5, fludarabine 30mg/m$^2$ days IV on days -6 through -2, and total body irradiation (200cGy or 400cGy) on day -1. All patients received post-transplant cyclophosphamide-based graft versus host disease prophylaxis consisting of cyclophosphamide at a dose of 50 mg/m$^2$ on days 3 and 4, mycophenolate mofetil from days 5 to 35, and tacrolimus or sirolimus from days 5 to 60, 90 or 180. Grafts were derived from peripheral blood or bone marrow. Pre-alloHCT samples were obtained at a median of 28 days prior to transplantation (range: 0-52 days). Post-alloHCT samples were obtained at a median of 57 days after transplantation (range: 21-405 days).

#### 2. Next generation Sequencing (NGS)

DNA extracted from baseline and relapse bone marrow samples were analyzed by targeted error-corrected next generation sequencing (NGS) using the 75-gene Archer VariantPlex myeloid panel utilizing anchored multiplex polymerase chain reaction (PCR) chemistry (ArcherDx Boulder, CO). The manufacturer's instructions were followed to generate sequencing-ready NGS libraries. In brief, 50ng of input DNA, diluted in 50uL of 10mM Tris-HCl was enzymatically fragmented, phosphorylated, and A-tailed. This was followed by ligation of adapters containing the Illumina p5 index and unique molecular barcode sequences that allowed for deduplication and error correction during bioinformatics analysis. Finally, two rounds of nested PCR amplification using genespecific primers (GSP1 and 2) were carried out to enrich targets of interest and introduce the Illumina p7 index sequence into the library.

For the baseline samples, liquid handling robots were used for library preparation for 48 samples. The remaining 8 baseline and all relapse samples were prepared manually. When robots were used, all pre-PCR steps were carried out on the Sciclone G3 NGS Workstation (PerkinElmer Health Sciences, Inc., Shelton, CT) while the post-PCR steps were performed on the Zephyr G3 NGS Workstation (PerkinElmer Health Sciences, Inc., Shelton, CT).

Libraries were subjected to paired-end 150bp sequencing utilizing unique dual indices on the NovaSeq 6000 system (Illumina, San Diego, CA), according to the manufacturer's instructions. Ten million reads were targeted per library.

A smaller, custom panel targeting recurrent hotspot mutations in the *FLT3*, *IDH1*, *IDH2*, *KIT,* and *NPM1* genes (VariantPlex, ArcherDx Boulder, CO) was used to identify persistent *IDH* mutations in a subset of pre- and post-alloHCT samples that had insufficient DNA concentrations for digital PCR reactions or rare mutations for which a targeted assay was unavailable. Libraries were prepared and sequenced as previously described (1).

Raw sequencing FASTQ files are available on the NCBI Sequence Reads Archive (SRA) database (PRJNA998386).

#### 3. Bioinformatics analysis

Demultiplexed FASTQ files were analyzed using the Archer Analysis software version 6.2.7 (ArcherDx Inc., Boulder, CO). Single nucleotide variant (SNV) and DNA structural variation (SV) pipelines were used. In short, raw reads were filtered based on quality scores outputted by the sequencing machine. De-duplication and error correction were performed by using unique molecular identifiers to obtain a single consensus read from each read family. At least 3 reads per barcode were used to define a 'deep-bin'. Reads were mapped to human genome build GRCh37 using the Bowtie2 and BWA-Mem aligners. Variant calling was performed using the Freebayes and LoFreq callers using a minimum allele frequency threshold of 0.001. Variants were annotated using Variant Effect Predictor.

To compensate for background error, libraries prepared from Genome in a Bottle DNA (Coriell Institute for Medical Research, Camden, NJ) were used to develop a Normal dataset (ND). The ND-background error model was generated for each variant and used to calculate a p-value at each nucleotide position.

For variants in the diagnostic or relapse samples, the following filters were applied to identify relevant variants as follows:

1. Alternate Observations ≥ 5
2. Unique Start for Alternate Observations ≥ 3
3. VEP consequence = coding_sequence_variant, feature_elongation, feature_truncation, frameshift_variant, incomplete_terminal_codon_variant, inframe_deletion, inframe_insertion, missense_variant, protein_altering_variant, splice_acceptor_variant, splice_donor_variant, splice_region_variant, start_lost, stop_gained, stop_lost, transcript_ablation, transcript_amplification
4. Allele Fraction ≥ 0.01
5. gnomAD Allele Frequency ≤ 0.001
6. Has Sample Strand Bias = No
7. Has Sequencing Direction Bias = No
8. Deep Alternate Observations ≥ 5
9. ND DAF Outlier p-value ≤ 0.0001
10. Homopolymer run ≤ 9

For variants in the remission samples, filters were applied as previously described (1).

### 4. Digital Droplet Polymerase Chain Reaction (ddPCR)

Assay setup for all ddPCR reactions was carried out according to manufacturer's recommendations (Bio-Rad Laboratories, Inc., Hercules, CA). All assays used mutant and wild-type specific probes tagged with FAM and HEX fluorophores, respectively. Positive, negative, and no-template controls were included in each run.

300-330ng of input DNA was used per 22uL reaction, which was combined with 1X ddPCR Supermix for probes (no dUTP) (cat # 1863023, Bio-Rad Laboratories, Inc., Hercules, CA), 1X ddPCR mutation detection primers/probes (full list provided in **Supplementary Table 2**) and 5U MsEI restriction enzyme (cat # R1040S, New England BioLabs, Inc., Ipswich, MA). The plate was incubated at room temperature for 3 minutes. 20uL of each reaction was combined with 70uL of Automated Droplet Generation Oil for Probes (cat # 1864110). The resulting mix was used to generate ~20,000 droplets on the Automated Droplet Generator (Bio-Rad Laboratories, Inc., Hercules, CA). PCR was carried out according to the protocol supplied by the manufacturer in a thermocycler using a heated lid at 105°C and a ramp rate of 2°C per second. An initial denaturation step at 95°C for 10 minutes was followed by 40 cycles each of 94°C for 30 seconds and 55°C for 1 minute with a final enzyme deactivation step of 98°C for 1 minute. Fluorescence signals from individual droplets were analyzed on the QX200 Droplet Reader (cat # 18640003, Bio-Rad Laboratories, Inc., Hercules, CA). Data was generated as .qlp files which were assessed using the QuantaSoft Analysis Pro ver. 1.0.596 software (Bio-Rad Laboratories, Inc., Hercules, CA). After ensuring an adequate number of droplets in each well (>10,000), thresholds were applied using negative controls to exclude false positive droplets from analysis. Recommendations from the manufacturer were followed to confidently identify positive events (2).

Each assay was previously validated by using a known positive control DNA sample which was mixed with healthy donor DNA to obtain a dilution series from 5% - 0.01% variant allele frequency. ddPCR reactions were set up and analyzed at each dilution point using the protocol outlined above to determine the limit of detection (LOD) for individual assays (see **Supplementary Table 3**).

5. **Statistical analysis**

Clinical endpoints of overall survival (OS) and relapse were evaluated with the day of transplant as day 0. Comparison between groups was performed using the Mann Whitney U test, Chi-Square test or Fisher's Exact test on GraphPad Prism ver 9.3.1. Kaplan-Meier estimation and log-rank tests were used for OS. Fine-Gray regression model was used for the cumulative incidence of relapse with non-relapse mortality as a competing risk. Figures were created using R version 4.2.1.

**Supplementary Figure 1:** Schematic representation of patients included/samples analyzed in the study. AML, acute myeloid leukemia; ddPCR, digital droplet PCR; ecNGS, error-corrected next generation sequencing.

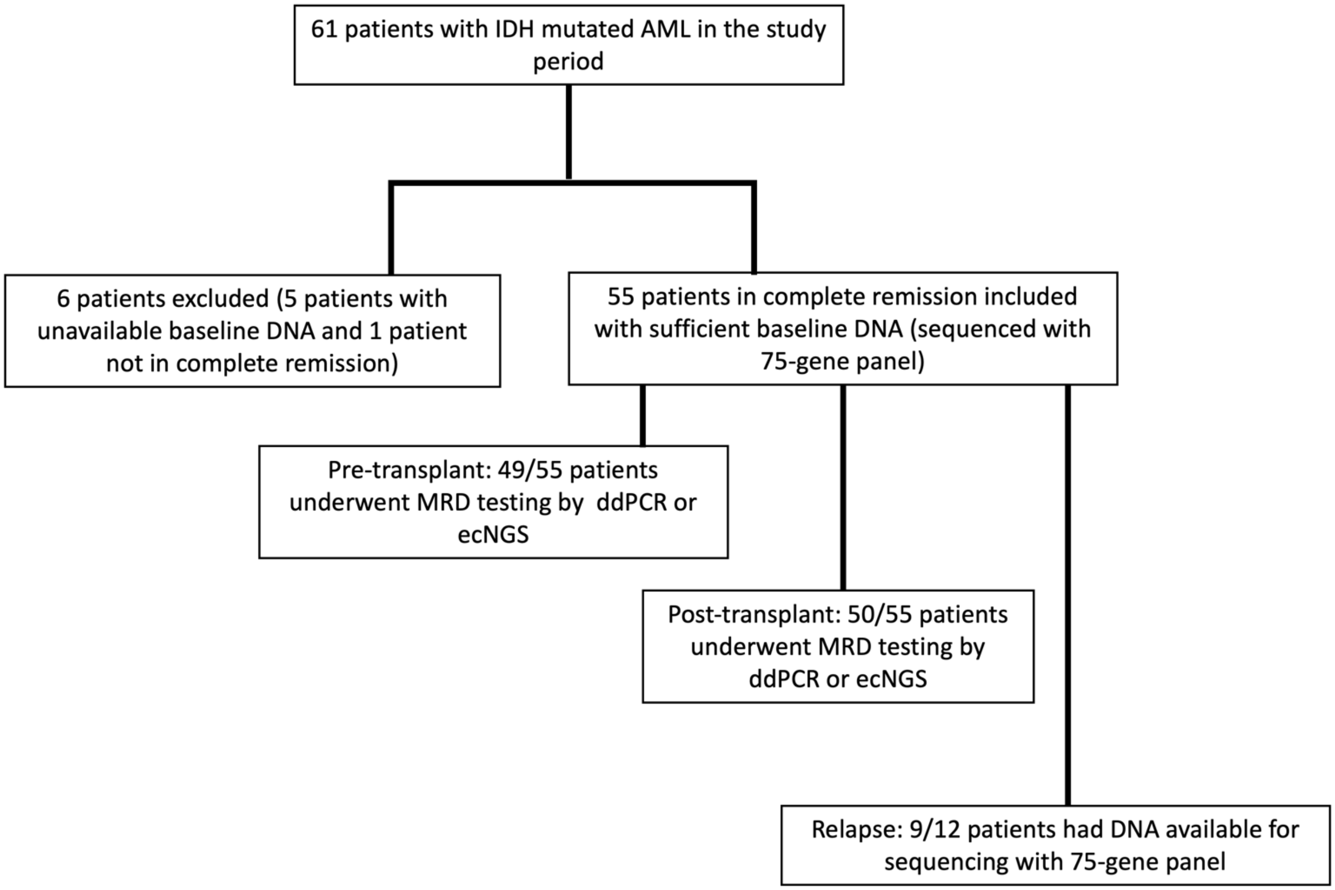

**Supplementary Figure 2:**

Co-mutational spectrum of *IDH*-mutated patients._alloHCT, allogeneic hematopoietic cell transplant

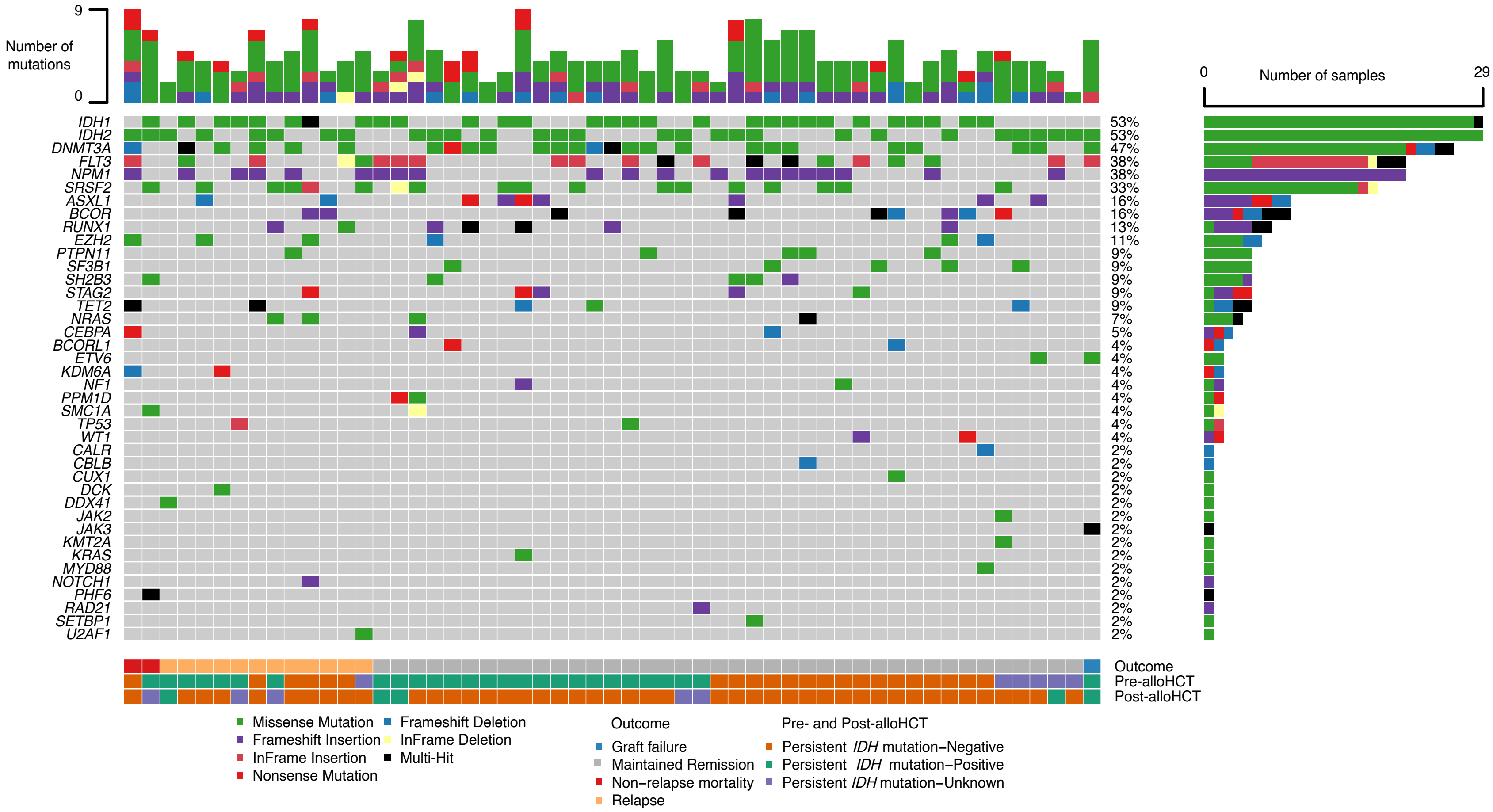

**Supplementary Table 1:**

Patient Clinical Characteristics. ELN, European Leukemia Net

| Parameter | *IDH1* (n=29) | *IDH2* (n=29) |
|---|---|---|
| **Age median (range)** | 62.9 (27.8-74.9) | 63.3 (43.1-74.4) |
| **Sex, n (%)** | | |
| Female | 13 (44.8) | 16 (55.2) |
| Male | 16 (55.2) | 13 (44.8) |
| **Cytogenetics, n (%)** | | |
| Favorable | 1 (3.4) | 0 (0) |
| Intermediate | 21 (72.4) | 23 (79.4) |
| Adverse | 3 (10.4) | 3 (10.3) |
| Unknown | 4 (13.8) | 3 (10.3) |
| **ELN risk category, n (%)** | | |
| Favorable | 18 (62.1) | 6 (20.7) |
| Intermediate | 5 (17.2) | 17 (58.6) |
| Adverse | 6 (20.7) | 6 (20.7) |
| **IDH inhibitor, n (%)** | | |
| Pre-Transplant | 6 (20) | 11 (38) |
| Post-Transplant | 5 (17) | 8 (28) |
| **Remission number, n (%)** | | |
| 1 | 26 (89.6) | 29 (100) |
| 2 | 3 (10.4) | 0 (0) |
| **Type of transplant, n (%)** | | |
| Haploidentical | 24 (82.7) | 27 (93.2) |
| Matched unrelated | 1 (3.4) | 1 (3.4) |
| Mismatched unrelated | 4 (13.8) | 1 (3.4) |
| **Pre-Transplant Persistent IDH mutation, n (%)** | | |
| Positive | 14 (48.3) | 14 (48.3) |
| Negative | 14 (48.3) | 10 (34.5) |
| Not performed | 1 (3.4) | 5 (17.2) |
| **Post-Transplant Persistent IDH mutation, n (%)** | | |
| Positive | 2 (6.6) | 3 (10.3) |
| Negative | 24 (80) | 23 (79.4) |
| Not performed | 3 (10.4) | 3 (10.3) |

**Supplementary Table 2:**

Persistent *IDH* mutation status and relapse outcomes in patients with detectable measurable residual disease by flow cytometry. MRD, Measurable Residual Disease; *IDH*m, persistent *IDH* mutation; alloHCT, allogeneic hematopoietic cell transplant

| Patient number | MRD status by Flow cytometry | *IDH*m status pre-alloHCT | Relapse |
|---|---|---|---|
| 3 | Positive | Positive | Yes |
| 7 | Positive | Negative | Yes |
| 15 | Positive | Positive | No |
| 41 | Positive | Positive | No |
| 43 | Positive | Unknown | No |

**Supplementary Table 3**:

Bio-Rad ddPCR assays used with their limit of detection (LOD).

| Mutation | Assay ID | LOD determined by dilution experiments |
|---|---|---|
| *IDH1* R132H | dHsaMDV2010055 | 0.05% |
| *IDH1* R132C | dHsaMDV2010053 | 0.05% |
| *IDH1* R132G | dHsaMDV2510512 | 0.05% |
| *IDH2* R140W | dHsaIS2502317, dHsaIS2502316 | 0.05% |
| *IDH2* R140Q | dHsaMDV2010057 | 0.05% |
| *IDH2* R140L | dHsaMDS2515782 | 0.05% |
| *IDH2* R172K | dHsaMDV2010059 | 0.01% |

**Supplementary Table 4:**

List of variants identified at baseline in *IDH*-mutated AML patients, persistent *IDH* mutation status pre- and post-transplant and variants present at relapse. alloHCT, allogeneic hematopoietic cell transplant; NA, Not Available.

| | Baseline | | | | Pre-alloHCT Persistent IDH mutation | Post-alloHCT Persistent IDH mutation | Relapse | | | |
|---|---|---|---|---|---|---|---|---|---|---|
| **Patient number** | **Genes mutated** | **Protein change** | **Nucleotide change** | **Allelic Fraction** | **Allelic Fraction** | **Allelic Fraction** | **Genes mutated** | **Protein change** | **Nucleotide change** | **Allelic Fraction** |
| 1 | IDH1 | NP_005887.2:p.Arg132Cys | NM_005896.3:c.394C>T | 0.4376 | 0.041 | NA | IDH1 | NP_005887.2:p.Arg132Cys | NM_005896.3:c.394C>T | 0.3706 |
| | NPM1 | NP_002511.1:p.Trp288CysfsTer12 | NM_002520.6:c.863_864insCAGA | 0.4319 | | | TP53 | NP_000537.3:p.Pro219Ser | NM_000546.5:c.655C>T | 0.0479 |
| | TP53 | NP_000537.3:p.Arg280dup | NM_000546.5:c.838_840dup | 0.1702 | | | | | | |
| 2 | IDH1 | NP_005887.2:p.Arg132His | NM_005896.3:c.395G>A | 0.2157 | 0 | 0 | IDH1 | NP_005887.2:p.Arg132His | NM_005896.3:c.395G>A | 0.0258 |
| | IDH2 | NP_002159.2:p.Arg140Trp | NM_002168.3:c.418C>T | 0.0115 | 0 | 0 | ASXL1 | NP_056153.2:p.Gly646TrpfsTer12 | NM_015338.5:c.1934dup | 0.0614 |
| | DNMT3A | NP_072046.2:p.Arg882His | NM_022552.4:c.2645G>A | 0.4303 | | | DNMT3A | NP_072046.2:p.Arg882His | NM_022552.4:c.2645G>A | 0.0238 |
| | FLT3 | NP_004110.2:p.Asp593_Leu601dup | NM_004119.2:c.1776_1802dup | 0.0907 | | | PTPN11 | NP_002825.3:p.Gln510Leu | NM_002834.4:c.1529A>T | 0.0217 |
| | NPM1 | NP_002511.1:p.Trp288CysfsTer12 | NM_002520.6:c.860_863dup | 0.1711 | | | TET2 | NP_001120680.1:p.Arg1516Ter | NM_001127208.2:c.4546C>T | 0.0245 |
| | TET2 | NP_060098.3:p.Thr229AsnfsTer25 | NM_017628.4:c.685dup | 0.1404 | | | NPM1 | NP_002511.1:p.Trp288CysfsTer12 | NM_002520.6:c.860_863dup | 0.0071 |
| | TET2 | NP_001120680.1:p.Arg1516Ter | NM_001127208.2:c.4546C>T | 0.4234 | | | | | | |
| 3 | IDH2 | NP_002159.2:p.Arg140Trp | NM_002168.3:c.418C>T | 0.473 | 0.0022 | NA | IDH2 | NP_002159.2:p.Arg140Trp | NM_002168.3:c.418C>T | 0.3562 |
| | NRAS | NP_002515.1:p.Gly12Asp | NM_002524.4:c.35G>A | 0.4411 | | | NRAS | NP_002515.1:p.Gly12Asp | NM_002524.4:c.35G>A | 0.3649 |
| | ASXL1 | NP_056153.2:p.Gly646TrpfsTer12 | NM_015338.5:c.1934dup | 0.0376 | | | RUNX1 | NP_001745.2:p.Ala251CysfsTer10 | NM_001754.4:c.749_750insC | 0.0706 |
| | RUNX1 | NP_001745.2:p.Ala251CysfsTer10 | NM_001754.4:c.749_750insC | 0.1037 | | | SRSF2 | NP_003007.2:p.Pro95Thr | NM_003016.4:c.283C>A | 0.3486 |
| | SRSF2 | NP_003007.2:p.Pro95Thr | NM_003016.4:c.283C>A | 0.4506 | | | WT1 | NP_000369.3:p.Thr429TyrfsTer9 | NM_000378.4:c.1283_1284insC | 0.0569 |
| | | | | | | | ASXL1 | NP_056153.2:p.Gly646TrpfsTer12 | NM_015338.5:c.1934dup | 0.022 |
| 4 | IDH1 | NP_005887.2:p.Arg132Cys | NM_005896.3:c.394C>T | 0.4506 | 0 | 0 | | | | |
| | DNMT3A | NP_072046.2:p.Arg882Cys | NM_022552.4:c.2644C>T | 0.454 | | | | | | |
| | NPM1 | NP_002511.1:p.Trp288CysfsTer12 | NM_002520.6:c.860_863dup | 0.1747 | | | | | | |
| | PTPN11 | NP_002825.3:p.Ala72Thr | NM_002834.4:c.214G>A | 0.4549 | | | | | | |
| | SRSF2 | NP_003007.2:p.Pro95His | NM_003016.4:c.284C>A | 0.4734 | | | | | | |
| 5 | IDH1 | NP_005887.2:p.Arg132Cys | NM_005896.3:c.394C>T | 0.4699 | 0.0649 | 0 | IDH1 | NP_005887.2:p.Arg132Cys | NM_005896.3:c.394C>T | 0.4325 |
| | DNMT3A | NP_072046.2:p.Arg688Cys | NM_022552.4:c.2062C>T | 0.5036 | | | DNMT3A | NP_072046.2:p.Arg688Cys | NM_022552.4:c.2062C>T | 0.4085 |
| | DNMT3A | NP_072046.2:p.Arg598Ter | NM_022552.4:c.1792C>T | 0.5085 | | | DNMT3A | NP_072046.2:p.Arg598Ter | NM_022552.4:c.1792C>T | 0.4372 |
| | FLT3 | NP_004110.2:p.Asp835Tyr | NM_004119.2:c.2503G>T | 0.48 | | | FLT3 | NP_004110.2:p.Asp835Tyr | NM_004119.2:c.2503G>T | 0.4892 |
| | NPM1 | NP_002511.1:p.Trp288CysfsTer12 | NM_002520.6:c.860_863dup | 0.2521 | | | NPM1 | NP_002511.1:p.Trp288CysfsTer12 | NM_002520.6:c.860_863dup | 0.1736 |
| 6 | IDH1 | NP_005887.2:p.Arg132Leu | NM_005896.3:c.395G>T | 0.2169 | 0 | 0 | SRSF2 | NP_003007.2:p.Arg94dup | NM_003016.4:c.283_284insGGC | 0.1688 |
| | IDH1 | NP_005887.2:p.Arg132Cys | NM_005896.3:c.394C>T | 0.2019 | 0 | 0 | STAG2 | NP_006594.3:p.Arg110Ter | NM_006603.4:c.328C>T | 0.3658 |

| | Baseline | | | | Pre-alloHCT Persistent IDH mutation | Post-alloHCT Persistent IDH mutation | Relapse | | | |
|---|---|---|---|---|---|---|---|---|---|---|
| **Patient number** | **Genes mutated** | **Protein change** | **Nucleotide change** | **Allelic Fraction** | **Allelic Fraction** | **Allelic Fraction** | **Genes mutated** | **Protein change** | **Nucleotide change** | **Allelic Fraction** |
| 1 | IDH1 | NP_005887.2:p.Arg132Cys | NM_005896.3:c.394C>T | 0.4376 | 0.041 | NA | IDH1 | NP_005887.2:p.Arg132Cys | NM_005896.3:c.394C>T | 0.3706 |
| | NPM1 | NP_002511.1:p.Trp288CysfsTer12 | NM_002520.6:c.863_864insCAGA | 0.4319 | | | TP53 | NP_000537.3:p.Pro219Ser | NM_000546.5:c.655C>T | 0.0479 |
| | TP53 | NP_000537.3:p.Arg280dup | NM_000546.5:c.838_840dup | 0.1702 | | | | | | |
| | BCOR | NP_060215.4:p.Ala603GlyfsTer8 | NM_017745.5:c.1807dup | 0.9592 | | | TET2 | NP_060098.3:p.Gly342Ter | NM_017628.4:c.1024G>T | 0.1608 |
| | EZH2 | NP_004447.2:p.Glu745Lys | NM_004456.4:c.2233G>A | 0.0258 | | | | | | |
| | NRAS | NP_002515.1:p.Gly12Asp | NM_002524.4:c.35G>A | 0.4816 | | | | | | |
| | NOTCH1 | NP_060087.3:p.Gln2343AlafsTer11 | NM_017617.4:c.7025dup | 0.0122 | | | | | | |
| | SRSF2 | NP_003007.2:p.Arg94dup | NM_003016.4:c.283_284insGGC | 0.3909 | | | | | | |
| | STAG2 | NP_006594.3:p.Arg110Ter | NM_006603.4:c.328C>T | 0.96 | | | | | | |
| 7 | IDH2 | NP_002159.2:p.Arg140Gln | NM_002168.3:c.419G>A | 0.4941 | 0 | 0 | IDH2 | NP_002159.2:p.Arg140Gln | NM_002168.3:c.419G>A | 0.3928 |
| | ASXL1 | NP_056153.2:p.Pro808LeufsTer10 | NM_015338.5:c.2423del | 0.4771 | | | ASXL1 | NP_056153.2:p.Pro808LeufsTer10 | NM_015338.5:c.2423del | 0.3772 |
| | BCOR | NP_060215.4:p.Gly81GlufsTer12 | NM_017745.5:c.241_242insA | 0.967 | | | BCOR | NP_060215.4:p.Gly81GlufsTer12 | NM_017745.5:c.241_242insA | 0.6523 |
| | | | | | | | ETV6 | NP_001978.1:p.Asp46_Ser47insTer | NM_001987.4:c.139_140insGAT | 0.1804 |
| 8 | IDH2 | NP_002159.2:p.Arg172Lys | NM_002168.3:c.515G>A | 0.4742 | 0 | 0 | | | | |
| | NPM1 | NP_002511.1:p.Trp288CysfsTer12 | NM_002520.6:c.864delinsCCAGT | 0.402 | | | | | | |
| 9 | IDH2 | NP_002159.2:p.Arg140Gln | NM_002168.3:c.419G>A | 0.4731 | 0 | 0 | | | | |
| | ASXL1 | NP_056153.2:p.Tyr591Ter | NM_015338.5:c.1772dup | 0.4699 | | | | | | |
| | BCOR | NP_060215.4:p.Leu1545SerfsTer8 | NM_017745.5:c.4632_4633insAGCC | 0.2021 | | | | | | |
| | BCOR | NP_060215.4:p.Glu867Ter | NM_017745.5:c.2599G>T | 0.3517 | | | | | | |
| | BCOR | NP_060215.4:p.Arg1513Ter | NM_017745.5:c.4537C>T | 0.0528 | | | | | | |
| | SH2B3 | NP_005466.1:p.Val465Ile | NM_005475.2:c.1393G>A | 0.5227 | | | | | | |
| | SRSF2 | NP_003007.2:p.Pro95His | NM_003016.4:c.284C>A | 0.4584 | | | | | | |
| | STAG2 | NP_006594.3:p.Cys772MetfsTer13 | NM_006603.4:c.2313dup | 0.8842 | | | | | | |
| 10 | IDH2 | NP_002159.2:p.Arg140Gln | NM_002168.3:c.419G>A | 0.424 | 0.36 | 0 | | | | |
| | CEBPA | NP_004355.2:p.His24AlafsTer84 | NM_004364.4:c.68dup | 0.1187 | | | | | | |
| | FLT3 | NP_004110.2:p.Arg595_Leu601dup | NM_004119.2:c.1784_1804dup | 0.0455 | | | | | | |
| | NPM1 | NP_002511.1:p.Trp288CysfsTer12 | NM_002520.6:c.860_863dup | 0.2079 | | | | | | |

| | Baseline | | | | Pre-alloHCT Persistent IDH mutation | Post-alloHCT Persistent IDH mutation | Relapse | | | |
|---|---|---|---|---|---|---|---|---|---|---|
| **Patient number** | **Genes mutated** | **Protein change** | **Nucleotide change** | **Allelic Fraction** | **Allelic Fraction** | **Allelic Fraction** | **Genes mutated** | **Protein change** | **Nucleotide change** | **Allelic Fraction** |
| 1 | IDH1 | NP_005887.2:p.Arg132Cys | NM_005896.3:c.394C>T | 0.4376 | 0.041 | NA | IDH1 | NP_005887.2:p.Arg132Cys | NM_005896.3:c.394C>T | 0.3706 |
| | NPM1 | NP_002511.1:p.Trp288CysfsTer12 | NM_002520.6:c.863_864insCAGA | 0.4319 | | | TP53 | NP_000537.3:p.Pro219Ser | NM_000546.5:c.655C>T | 0.0479 |
| | TP53 | NP_000537.3:p.Arg280dup | NM_000546.5:c.838_840dup | 0.1702 | | | | | | |
| | NRAS | NP_002515.1:p.Gln61Leu | NM_002524.4:c.182A>T | 0.2598 | | | | | | |
| | PPM1D | NP_003611.1:p.Thr501Ala | NM_003620.3:c.1501A>G | 0.5011 | | | | | | |
| | SMC1A | NP_006297.2:p.Lys454del | NM_006306.3:c.1360_1362del | 0.2643 | | | | | | |
| | SRSF2 | NP_003007.2:p.Pro95Leu | NM_003016.4:c.284C>T | 0.5137 | | | | | | |
| 11 | IDH1 | NP_005887.2:p.Arg132Gly | NM_005896.3:c.394C>G | 0.4624 | 0.0068 | 0.0005 | | | | |
| | FLT3 | NP_004110.2:p.Thr582_Arg607dup | NM_004119.2:c.1743_1820dup | 0.0169 | | | | | | |
| | DNMT3A | | NM_022552.4:c.1015-1G>A | 0.4905 | | | | | | |
| | NPM1 | NP_002511.1:p.Trp288CysfsTer12 | NM_002520.6:c.860_863dup | 0.2186 | | | | | | |
| 12 | IDH2 | NP_002159.2:p.Arg140Gln | NM_002168.3:c.419G>A | 0.4522 | 0.0277 | 0 | | | | |
| | DNMT3A | NP_072046.2:p.Arg882His | NM_022552.4:c.2645G>A | 0.4268 | | | | | | |
| | EZH2 | NP_004447.2:p.Leu101Ter | NM_004456.4:c.301del | 0.6249 | | | | | | |
| | RUNX1 | NP_001745.2:p.Arg169LysfsTer44 | NM_001754.4:c.505dup | 0.8296 | | | | | | |
| | SH2B3 | NP_005466.1:p.Arg43Cys | NM_005475.2:c.127C>T | 0.4966 | | | | | | |
| 13 | IDH1 | NP_005887.2:p.Arg132Cys | NM_005896.3:c.394C>T | 0.3653 | 0 | 0 | | | | |
| | IDH2 | NP_002159.2:p.Arg140Gln | NM_002168.3:c.419G>A | 0.0639 | 0 | 0 | | | | |
| | DNMT3A | NP_072046.2:p.Arg882His | NM_022552.4:c.2645G>A | 0.4549 | | | | | | |
| | FLT3 | NP_004110.2:p.Asp835Glu | NM_004119.2:c.2505T>G | 0.3617 | | | | | | |
| | NPM1 | NP_002511.1:p.Trp288CysfsTer12 | NM_002520.6:c.863_864insCCTG | 0.3209 | | | | | | |
| | SETBP1 | NP_056374.2:p.Glu931Lys | NM_015559.2:c.2791G>A | 0.4888 | | | | | | |
| | SH2B3 | NP_005466.1:p.Ser213Arg | NM_005475.2:c.639C>A | 0.4801 | | | | | | |
| | FLT3 | ITD | | 0.001 | | | | | | |
| 14 | IDH1 | NP_005887.2:p.Arg132His | NM_005896.3:c.395G>A | 0.3995 | 0 | 0 | | | | |
| | CEBPA | NP_004355.2:p.Pro204ArgfsTer114 | NM_004364.4:c.611del | 0.3907 | | | | | | |
| | DNMT3A | NP_072046.2:p.Arg556Ser | NM_022552.4:c.1668G>T | 0.0178 | | | | | | |

| | Baseline | | | | Pre-alloHCT Persistent IDH mutation | Post-alloHCT Persistent IDH mutation | Relapse | | | |
|---|---|---|---|---|---|---|---|---|---|---|
| **Patient number** | **Genes mutated** | **Protein change** | **Nucleotide change** | **Allelic Fraction** | **Allelic Fraction** | **Allelic Fraction** | **Genes mutated** | **Protein change** | **Nucleotide change** | **Allelic Fraction** |
| 1 | IDH1 | NP_005887.2:p.Arg132Cys | NM_005896.3:c.394C>T | 0.4376 | 0.041 | NA | IDH1 | NP_005887.2:p.Arg132Cys | NM_005896.3:c.394C>T | 0.3706 |
| | NPM1 | NP_002511.1:p.Trp288CysfsTer12 | NM_002520.6:c.863_864insCAGA | 0.4319 | | | TP53 | NP_000537.3:p.Pro219Ser | NM_000546.5:c.655C>T | 0.0479 |
| | TP53 | NP_000537.3:p.Arg280dup | NM_000546.5:c.838_840dup | 0.1702 | | | | | | |
| | NPM1 | NP_002511.1:p.Trp288CysfsTer12 | NM_002520.6:c.863_864insCCTG | 0.2234 | | | | | | |
| | SF3B1 | NP_036565.2:p.Tyr765Cys | NM_012433.3:c.2294A>G | 0.0333 | | | | | | |
| | SRSF2 | NP_003007.2:p.Pro95Arg | NM_003016.4:c.284C>G | 0.4017 | | | | | | |
| 15 | IDH2 | NP_002159.2:p.Arg140Gln | NM_002168.3:c.419G>A | 0.3855 | 0.0759 | 0 | | | | |
| | BCOR | | NM_017745.5:c.4874+3G>C | 0.5974 | | | | | | |
| | BCOR | | NM_017745.5:c.4639+1G>C | 0.0277 | | | | | | |
| | BCOR | | NM_017745.5:c.4326+1G>C | 0.0667 | | | | | | |
| | BCORL1 | NP_068765.3:p.Gln1341Ter | NM_021946.4:c.4021C>T | 0.0155 | | | | | | |
| | DNMT3A | NP_072046.2:p.Tyr584Ter | NM_022552.4:c.1752C>A | 0.7533 | | | | | | |
| | SF3B1 | NP_036565.2:p.Arg625Cys | NM_012433.3:c.1873C>T | 0.3949 | | | | | | |
| 16 | IDH2 | NP_002159.2:p.Arg140Gln | NM_002168.3:c.419G>A | 0.4865 | 0.47 | NA | | | | |
| | DNMT3A | NP_072046.2:p.Arg882Pro | NM_022552.4:c.2645G>C | 0.4963 | | | | | | |
| | SRSF2 | NP_003007.2:p.Pro95Leu | NM_003016.4:c.284C>T | 0.5159 | | | | | | |
| 17 | IDH1 | NP_005887.2:p.Arg132Gly | NM_005896.3:c.394C>G | 0.4958 | 0.0066 | NA | | | | |
| | DNMT3A | NP_072046.2:p.Glu774= | NM_022552.4:c.2322G>A | 0.4912 | | | | | | |
| | FLT3 | NP_004110.2:p.Val592_Tyr599dup | NM_004119.2:c.1773_1796dup | 0.3131 | | | | | | |
| | RAD21 | NP_006256.1:p.Cys392LeufsTer11 | NM_006265.2:c.1174dup | 0.4807 | | | | | | |
| 18 | IDH2 | NP_002159.2:p.Arg140Trp | NM_002168.3:c.418C>T | 0.0518 | 0 | 0 | | | | |
| | DNMT3A | NP_072046.2:p.Gly421AlafsTer2 | NM_022552.4:c.1262_1266delinsCCTA | 0.4459 | | | | | | |
| | CEBPA | NP_004355.2:p.Gln221Ter | NM_004364.4:c.661C>T | 0.1886 | | | | | | |
| | EZH2 | NP_004447.2:p.Arg690His | NM_004456.4:c.2069G>A | 0.1472 | | | | | | |
| | FLT3 | NP_004110.2:p.Phe590_Asp600dup | NM_004119.2:c.1767_1799dup | 0.0434 | | | | | | |
| | KDM6A | NP_066963.2:p.Gly681AspfsTer10 | NM_021140.3:c.2042del | 0.0233 | | | | | | |
| | NPM1 | NP_002511.1:p.Trp288CysfsTer12 | NM_002520.6:c.860_863dup | 0.2236 | | | | | | |

| | Baseline | | | | Pre-alloHCT Persistent IDH mutation | Post-alloHCT Persistent IDH mutation | Relapse | | | |
|---|---|---|---|---|---|---|---|---|---|---|
| Patient number | Genes mutated | Protein change | Nucleotide change | Allelic Fraction | Allelic Fraction | Allelic Fraction | Genes mutated | Protein change | Nucleotide change | Allelic Fraction |
| 1 | IDH1 | NP_005887.2:p.Arg132Cys | NM_005896.3:c.394C>T | 0.4376 | 0.041 | NA | IDH1 | NP_005887.2:p.Arg132Cys | NM_005896.3:c.394C>T | 0.3706 |
| | NPM1 | NP_002511.1:p.Trp288CysfsTer12 | NM_002520.6:c.863_864insCAGA | 0.4319 | | | TP53 | NP_000537.3:p.Pro219Ser | NM_000546.5:c.655C>T | 0.0479 |
| | TP53 | NP_000537.3:p.Arg280dup | NM_000546.5:c.838_840dup | 0.1702 | | | | | | |
| | TET2 | NP_001120680.1:p.Gly1860Arg | NM_001127208.2:c.5578G>A | 0.4537 | | | | | | |
| | TET2 | NP_060098.3:p.Ser327Ter | NM_017628.4:c.980C>G | 0.4364 | | | | | | |
| 19 | IDH1 | NP_005887.2:p.Arg132His | NM_005896.3:c.395G>A | 0.4601 | 0.364 | 0.0013 | | | | |
| | PPM1D | NP_003611.1:p.Arg552Ter | NM_003620.3:c.1654C>T | 0.0152 | | | | | | |
| | NPM1 | NP_002511.1:p.Trp288CysfsTer12 | NM_002520.6:c.860_863dup | 0.2028 | | | | | | |
| | SRSF2 | NP_003007.2:p.Pro95_Arg117del | NM_003016.4:c.279_347del | 0.2848 | | | | | | |
| | FLT3 | ITD | | 0.002 | | | | | | |
| 20 | IDH1 | NP_005887.2:p.Arg132His | NM_005896.3:c.395G>A | 0.4387 | 0 | 0 | | | | |
| | DNMT3A | NP_072046.2:p.Gly543Asp | NM_022552.4:c.1628G>A | 0.4369 | | | | | | |
| | FLT3 | NP_004110.2:p.Phe453Leu | NM_004119.2:c.1359C>A | 0.2689 | | | | | | |
| | FLT3 | NP_004110.2:p.Lys623Ile | NM_004119.2:c.1868A>T | 0.0152 | | | | | | |
| | NPM1 | NP_002511.1:p.Trp288CysfsTer12 | NM_002520.6:c.860_863dup | 0.1851 | | | | | | |
| | PTPN11 | NP_002825.3:p.Gly60Val | NM_002834.4:c.179G>T | 0.0587 | | | | | | |
| | SH2B3 | NP_005466.1:p.Lys180TrpfsTer19 | NM_005475.2:c.533_537dup | 0.3914 | | | | | | |
| 21 | IDH1 | NP_005887.2:p.Arg132His | NM_005896.3:c.395G>A | 0.0896 | 0 | 0 | | | | |
| | CBLB | NP_733762.2:p.Pro428GlnfsTer20 | NM_170662.4:c.1283del | 0.4517 | | | | | | |
| | NPM1 | NP_002511.1:p.Trp288CysfsTer12 | NM_002520.6:c.860_863dup | 0.1662 | | | | | | |
| | NRAS | NP_002515.1:p.Gly13Asp | NM_002524.4:c.38G>A | 0.3458 | | | | | | |
| | NRAS | NP_002515.1:p.Gly13Cys | NM_002524.4:c.37G>T | 0.0254 | | | | | | |
| | NRAS | NP_002515.1:p.Gly12Ala | NM_002524.4:c.35G>C | 0.034 | | | | | | |
| | PTPN11 | NP_002825.3:p.Glu69Val | NM_002834.4:c.206A>T | 0.01 | | | | | | |
| 22 | IDH2 | NP_002159.2:p.Arg140Gln | NM_002168.3:c.419G>A | 0.43 | NA | 0 | | | | |
| | BCOR | NP_060215.4:p.Arg1163Ter | NM_017745.5:c.3487C>T | 0.025 | | | | | | |
| | JAK2 | NP_004963.1:p.Arg922Trp | NM_004972.3:c.2764C>T | 0.5415 | | | | | | |

| | Baseline | | | | Pre-alloHCT Persistent IDH mutation | Post-alloHCT Persistent IDH mutation | Relapse | | | |
|---|---|---|---|---|---|---|---|---|---|---|
| Patient number | Genes mutated | Protein change | Nucleotide change | Allelic Fraction | Allelic Fraction | Allelic Fraction | Genes mutated | Protein change | Nucleotide change | Allelic Fraction |
| 1 | IDH1 | NP_005887.2:p.Arg132Cys | NM_005896.3:c.394C>T | 0.4376 | 0.041 | NA | IDH1 | NP_005887.2:p.Arg132Cys | NM_005896.3:c.394C>T | 0.3706 |
| | NPM1 | NP_002511.1:p.Trp288CysfsTer12 | NM_002520.6:c.863_864insCAGA | 0.4319 | | | TP53 | NP_000537.3:p.Pro219Ser | NM_000546.5:c.655C>T | 0.0479 |
| | TP53 | NP_000537.3:p.Arg280dup | NM_000546.5:c.838_840dup | 0.1702 | | | | | | |
| | KMT2A | NP_005924.2:p.Glu2002Lys | NM_005933.3:c.6004G>A | 0.4835 | | | | | | |
| | SRSF2 | NP_003007.2:p.Pro95Arg | NM_003016.4:c.284C>G | 0.4099 | | | | | | |
| 23 | IDH1 | NP_005887.2:p.Arg132Cys | NM_005896.3:c.394C>T | 0.4487 | 0.33 | 0 | | | | |
| | DNMT3A | NP_072046.2:p.Arg882His | NM_022552.4:c.2645G>A | 0.4732 | | | | | | |
| | ASXL1 | NP_056153.2:p.Arg693Ter | NM_015338.5:c.2077C>T | 0.4923 | | | | | | |
| | RUNX1 | NP_001745.2:p.Ile364ValfsTer231 | NM_001754.4:c.1090_1103del | 0.0172 | | | | | | |
| | RUNX1 | NP_001745.2:p.Arg166Ter | NM_001754.4:c.496C>T | 0.0221 | | | | | | |
| 24 | IDH2 | NP_002159.2:p.Arg140Gln | NM_002168.3:c.419G>A | 0.0068 | 0.0049 | 0 | | | | |
| | DNMT3A | NP_072046.2:p.Arg736His | NM_022552.4:c.2207G>A | 0.1873 | | | | | | |
| 25 | IDH1 | NP_005887.2:p.Arg132His | NM_005896.3:c.395G>A | 0.4551 | 0 | 0 | | | | |
| | FLT3 | NP_004110.2:p.Asp835Tyr | NM_004119.2:c.2503G>T | 0.0875 | | | | | | |
| | NPM1 | NP_002511.1:p.Trp288CysfsTer12 | NM_002520.6:c.860_863dup | 0.2869 | | | | | | |
| | SRSF2 | NP_003007.2:p.Pro95Arg | NM_003016.4:c.284C>G | 0.4915 | | | | | | |
| 26 | IDH1 | NP_005887.2:p.Arg132Cys | NM_005896.3:c.394C>T | 0.4199 | 0.0016 | 0 | | | | |
| | ASXL1 | NP_056153.2:p.Gly646TrpfsTer12 | NM_015338.5:c.1934dup | 0.3632 | | | | | | |
| | SRSF2 | NP_003007.2:p.Pro95Arg | NM_003016.4:c.284C>G | 0.4466 | | | | | | |
| 27 | IDH2 | NP_002159.2:p.Arg140Gln | NM_002168.3:c.419G>A | 0.4513 | 0 | 0 | | | | |
| | NF1 | NP_000258.1:p.Lys77Asn | NM_000267.3:c.231A>T | 0.52 | | | | | | |
| | NPM1 | NP_002511.1:p.Trp288CysfsTer12 | NM_002520.6:c.860_863dup | 0.1675 | | | | | | |
| | SRSF2 | NP_003007.2:p.Pro95His | NM_003016.4:c.284C>A | 0.4627 | | | | | | |
| 28 | IDH1 | NP_005887.2:p.Arg132His | NM_005896.3:c.395G>A | 0.4385 | NA | 0 | | | | |
| | FLT3 | NP_004110.2:p.Gly846Ser | NM_004119.2:c.2536G>A | 0.0525 | | | | | | |
| | NPM1 | NP_002511.1:p.Trp288CysfsTer12 | NM_002520.6:c.860_863dup | 0.1879 | | | | | | |
| | SRSF2 | NP_003007.2:p.Pro95His | NM_003016.4:c.284C>A | 0.4444 | | | | | | |

| | Baseline | | | | Pre-alloHCT Persistent IDH mutation | Post-alloHCT Persistent IDH mutation | Relapse | | | |
|---|---|---|---|---|---|---|---|---|---|---|
| Patient number | Genes mutated | Protein change | Nucleotide change | Allelic Fraction | Allelic Fraction | Allelic Fraction | Genes mutated | Protein change | Nucleotide change | Allelic Fraction |
| 1 | IDH1 | NP_005887.2:p.Arg132Cys | NM_005896.3:c.394C>T | 0.4376 | 0.041 | NA | IDH1 | NP_005887.2:p.Arg132Cys | NM_005896.3:c.394C>T | 0.3706 |
| | NPM1 | NP_002511.1:p.Trp288CysfsTer12 | NM_002520.6:c.863_864insCAGA | 0.4319 | | | TP53 | NP_000537.3:p.Pro219Ser | NM_000546.5:c.655C>T | 0.0479 |
| | TP53 | NP_000537.3:p.Arg280dup | NM_000546.5:c.838_840dup | 0.1702 | | | | | | |
| | U2AF1 | NP_006749.1:p.Gln157Pro | NM_006758.2:c.470A>C | 0.0115 | | | | | | |
| 29 | IDH2 | NP_002159.2:p.Arg140Gln | NM_002168.3:c.419G>A | 0.481 | 0.1683 | 0 | | | | |
| | ASXL1 | NP_056153.2:p.Glu719ThrfsTer5 | NM_015338.5:c.2154_2157del | 0.4745 | | | | | | |
| | EZH2 | NP_004447.2:p.Cys535Ser | NM_004456.4:c.1603T>A | 0.4754 | | | | | | |
| | SRSF2 | NP_003007.2:p.Pro95His | NM_003016.4:c.284C>A | 0.4646 | | | | | | |
| | CBL | NP_005179.2:p.Arg149Ter | NM_005188.3:c.445C>T | 0.0105 | | | | | | |
| 30 | IDH2 | NP_002159.2:p.Arg140Gln | NM_002168.3:c.419G>A | 0.2427 | 0.0028 | 0.0129 | | | | |
| | DDX41 | NP_057306.2:p.Asp73Glu | NM_016222.3:c.219T>G | 0.5103 | | | | | | |
| 31 | IDH2 | NP_002159.2:p.Arg140Gln | NM_002168.3:c.419G>A | 0.3374 | 0.0832 | 0.5897 | IDH2 | NP_002159.2:p.Arg140Gln | NM_002168.3:c.419G>A | 0.5738 |
| | DNMT3A | NP_072046.2:p.Met880Val | NM_022552.4:c.2638A>G | 0.3985 | | | DNMT3A | NP_072046.2:p.Met880Val | NM_022552.4:c.2638A>G | 0.4935 |
| | ETV6 | NP_001978.1:p.Arg369Gln | NM_001987.4:c.1106G>A | 0.1547 | | | ETV6 | NP_001978.1:p.Arg369Gln | NM_001987.4:c.1106G>A | 0.0922 |
| | FLT3 | NP_004110.2:p.Glu598_Tyr599insSerLeuTyrValAspPheArgGluTyrGlu | NM_004119.2:c.1795_1796insCCCTCTACGTTGATTTCAGAGAATATGAAT | 0.0293 | | | IKZF1 | NP_006051.1:p.His195Tyr | NM_006060.5:c.583C>T | 0.0833 |
| | JAK3 | NP_000206.2:p.Ala573Val | NM_000215.3:c.1718C>T | 0.0685 | | | NRAS | NP_002515.1:p.Gly13Cys | NM_002524.4:c.37G>T | 0.0228 |
| | JAK3 | NP_000206.2:p.Met511Ile | NM_000215.3:c.1533G>T | 0.0771 | | | RUNX1 | NP_001745.2:p.Arg346ProfsTer254 | NM_001754.4:c.1036dup | 0.1635 |
| | | | | | | | RUNX1 | NP_001745.2:p.Gly199Trp | NM_001754.4:c.595G>T | 0.0807 |
| | | | | | | | SF3B1 | NP_036565.2:p.Leu929Phe | NM_012433.3:c.2787G>T | 0.0355 |
| | | | | | | | WT1 | NP_000369.3:p.Ser208AlafsTer38 | NM_000378.4:c.622_626del | 0.0596 |
| | | | | | | | JAK3 | NP_000206.2:p.Ala573Val | NM_000215.3:c.1718C>T | 0.0005 |
| 33 | IDH2 | NP_002159.2:p.Arg172Lys | NM_002168.3:c.515G>A | 0.4339 | 0 | 0 | IDH2 | NP_002159.2:p.Arg172Lys | NM_002168.3:c.515G>A | 0.4877 |
| | DNMT3A | NP_072046.2:p.Arg882Cys | NM_022552.4:c.2644C>T | 0.4306 | | | DNMT3A | NP_072046.2:p.Arg882Cys | NM_022552.4:c.2644C>T | 0.4931 |
| | FLT3 | NP_004110.2:p.Gln575del | NM_004119.2:c.1724_1726del | 0.3919 | | | FLT3 | NP_004110.2:p.Lys663Arg | NM_004119.2:c.1988A>G | 0.3908 |
| | RUNX1 | NP_001745.2:p.Val186Asp | NM_001754.4:c.557T>A | 0.8707 | | | RUNX1 | NP_001745.2:p.Val186Asp | NM_001754.4:c.557T>A | 0.9831 |
| | | | | | | | EZH2 | p.Phe678Ser | NM_004456:c.2033T>C | 0.012 |

| | Baseline | | | | Pre-alloHCT Persistent IDH mutation | Post-alloHCT Persistent IDH mutation | Relapse | | | |
|---|---|---|---|---|---|---|---|---|---|---|
| Patient number | Genes mutated | Protein change | Nucleotide change | Allelic Fraction | Allelic Fraction | Allelic Fraction | Genes mutated | Protein change | Nucleotide change | Allelic Fraction |
| 1 | IDH1 | NP_005887.2:p.Arg132Cys | NM_005896.3:c.394C>T | 0.4376 | 0.041 | NA | IDH1 | NP_005887.2:p.Arg132Cys | NM_005896.3:c.394C>T | 0.3706 |
| | NPM1 | NP_002511.1:p.Trp288CysfsTer12 | NM_002520.6:c.863_864insCAGA | 0.4319 | | | TP53 | NP_000537.3:p.Pro219Ser | NM_000546.5:c.655C>T | 0.0479 |
| | TP53 | NP_000537.3:p.Arg280dup | NM_000546.5:c.838_840dup | 0.1702 | | | | | | |
| | | | | | | | CSF3R | NP_000751.1:p.Lys785GlnfsTer4 | NM_000760.3:c.2352dup | 0.4911 |
| | | | | | | | CSF3R | NP_000751.1:p.Leu636Gln | NM_000760.3:c.1907T>A | 0.0586 |
| 34 | IDH1 | NP_005887.2:p.Arg132Cys | NM_005896.3:c.394C>T | 0.1615 | 0.0015 | 0 | IDH1 | NP_005887.2:p.Arg132Cys | NM_005896.3:c.394C>T | 0.1129 |
| | DNMT3A | NP_072046.2:p.Gly302Cys | NM_022552.4:c.904G>T | 0.1792 | | | DNMT3A | NP_072046.2:p.Gly302Cys | NM_022552.4:c.904G>T | 0.0947 |
| | DCK | NP_000779.1:p.Leu71Pro | NM_000788.2:c.212T>C | 0.5094 | | | DCK | NP_000779.1:p.Leu71Pro | NM_000788.2:c.212T>C | 0.1248 |
| | KDM6A | NP_066963.2:p.Gln1039Ter | NM_021140.3:c.3115C>T | 0.0221 | | | | | | |
| 35 | IDH1 | NP_005887.2:p.Arg132Cys | NM_005896.3:c.394C>T | 0.1957 | 0.0048 | 0 | | | | |
| | KRAS | NP_004976.2:p.Gly13Cys | NM_004985.4:c.37G>T | 0.0181 | | | | | | |
| | ASXL1 | NP_056153.2:p.Arg693Ter | NM_015338.5:c.2077C>T | 0.2531 | | | | | | |
| | NF1 | | NM_000267.3:c.5547-2A>G | 0.2501 | | | | | | |
| | NF1 | NP_000258.1:p.Met739IlefsTer10 | NM_000267.3:c.2216_2217insAA | 0.0972 | | | | | | |
| | RUNX1 | NP_001745.2:p.Ser402IlefsTer198 | NM_001754.4:c.1203_1204insA | 0.1072 | | | | | | |
| | RUNX1 | NP_001745.2:p.Arg201Gln | NM_001754.4:c.602G>A | 0.0162 | | | | | | |
| | STAG2 | NP_006594.3:p.Arg1033Ter | NM_006603.4:c.3097C>T | 0.3121 | | | | | | |
| | SRSF2 | NP_003007.2:p.Pro95His | NM_003016.4:c.284C>A | 0.2641 | | | | | | |
| | TET2 | NP_001120680.1:p.Met1456SerfsTer2 | NM_001127208.2:c.4367del | 0.0136 | | | | | | |
| 36 | IDH1 | NP_005887.2:p.Arg132His | NM_005896.3:c.395G>A | 0.0357 | 0 | 0 | | | | |
| | STAG2 | NP_006594.3:p.Ala428Gly | NM_006603.4:c.1283C>G | 0.0804 | | | | | | |
| | WT1 | NP_000369.3:p.Val354CysfsTer14 | NM_000378.4:c.1059dup | 0.0536 | | | | | | |
| | FLT3 | ITD | | 0.012 | | | | | | |
| 37 | IDH2 | NP_002159.2:p.Arg140Gln | NM_002168.3:c.419G>A | 0.2654 | 0.131 | 0 | | | | |
| | ASXL1 | NP_056153.2:p.Gly646TrpfsTer12 | NM_015338.5:c.1934dup | 0.2646 | | | | | | |
| | DNMT3A | NP_072046.2:p.Arg882His | NM_022552.4:c.2645G>A | 0.2856 | | | | | | |
| | STAG2 | NP_006594.3:p.Tyr331Ter | NM_006603.4:c.992dup | 0.2812 | | | | | | |

| | Baseline | | | | Pre-alloHCT Persistent IDH mutation | Post-alloHCT Persistent IDH mutation | Relapse | | | |
|---|---|---|---|---|---|---|---|---|---|---|
| Patient number | Genes mutated | Protein change | Nucleotide change | Allelic Fraction | Allelic Fraction | Allelic Fraction | Genes mutated | Protein change | Nucleotide change | Allelic Fraction |
| 1 | IDH1 | NP_005887.2:p.Arg132Cys | NM_005896.3:c.394C>T | 0.4376 | 0.041 | NA | IDH1 | NP_005887.2:p.Arg132Cys | NM_005896.3:c.394C>T | 0.3706 |
| | NPM1 | NP_002511.1:p.Trp288CysfsTer12 | NM_002520.6:c.863_864insCAGA | 0.4319 | | | TP53 | NP_000537.3:p.Pro219Ser | NM_000546.5:c.655C>T | 0.0479 |
| | TP53 | NP_000537.3:p.Arg280dup | NM_000546.5:c.838_840dup | 0.1702 | | | | | | |
| 38 | IDH2 | NP_002159.2:p.Arg172Lys | NM_002168.3:c.515G>A | 0.2108 | 0.0078 | 0 | | | | |
| | BCOR | NP_060215.4:p.Asp438AlafsTer7 | NM_017745.5:c.1313_1317del | 0.1429 | | | | | | |
| | BCOR | NP_060215.4:p.Ser336LeufsTer45 | NM_017745.5:c.1005dup | 0.0784 | | | | | | |
| | DNMT3A | NP_072046.2:p.Gly543Val | NM_022552.4:c.1628G>T | 0.4119 | | | | | | |
| | FLT3 | ITD | | 0.67 | | | | | | |
| 39 | IDH2 | NP_002159.2:p.Arg140Gln | NM_002168.3:c.419G>A | 0.3901 | 0.0019 | 0 | | | | |
| | DNMT3A | NP_072046.2:p.Gly543Cys | NM_022552.4:c.1627G>T | 0.4078 | | | | | | |
| | SRSF2 | NP_003007.2:p.Pro95His | NM_003016.4:c.284C>A | 0.4053 | | | | | | |
| | FLT3 | ITD | | 0.017 | | | | | | |
| 40 | IDH2 | NP_002159.2:p.Arg172Lys | NM_002168.3:c.515G>A | 0.3065 | 0 | 0 | | | | |
| | BCOR | NP_060215.4:p.Pro731ThrfsTer9 | NM_017745.5:c.2190dup | 0.2457 | | | | | | |
| | STAG2 | | NM_006603.4:c.3053+2T>G | 0.6596 | | | | | | |
| | BCOR | NP_060215.4:p.Arg1163Ter | NM_017745.5:c.3487C>T | 0.3865 | | | | | | |
| | SF3B1 | NP_036565.2:p.Lys700Glu | NM_012433.3:c.2098A>G | 0.3144 | | | | | | |
| 41 | IDH1 | NP_005887.2:p.Arg132Gly | NM_005896.3:c.394C>G | 0.3935 | 0.4546 | 0 | | | | |
| | DNMT3A | NP_072046.2:p.Arg604GlyfsTer47 | NM_022552.4:c.1810del | 0.3715 | | | | | | |
| | TET2 | NP_060098.3:p.Ser145Asn | NM_017628.4:c.434G>A | 0.5035 | | | | | | |
| | NPM1 | NP_002511.1:p.Trp288CysfsTer12 | NM_002520.6:c.860_863dup | 0.1281 | | | | | | |
| 42 | IDH1 | NP_005887.2:p.Arg132Cys | NM_005896.3:c.394C>T | 0.2937 | 0.1968 | 0 | | | | |
| | DNMT3A | NP_072046.2:p.Met513AsnfsTer33 | NM_022552.4:c.1537dup | 0.0706 | | | | | | |
| | RUNX1 | NP_001745.2:p.Gly394ArgfsTer206 | NM_001754.4:c.1178dup | 0.0675 | | | | | | |
| | DNMT3A | NP_072046.2:p.Gly543Cys | NM_022552.4:c.1627G>T | 0.3606 | | | | | | |
| 43 | IDH2 | NP_002159.2:p.Arg172Lys | NM_002168.3:c.515G>A | 0.315 | NA | 0 | | | | |
| | DNMT3A | NP_072046.2:p.Ala741Glu | NM_022552.4:c.2222C>A | 0.2937 | | | | | | |

| | Baseline | | | | Pre-alloHCT Persistent IDH mutation | Post-alloHCT Persistent IDH mutation | Relapse | | | |
|---|---|---|---|---|---|---|---|---|---|---|
| Patient number | Genes mutated | Protein change | Nucleotide change | Allelic Fraction | Allelic Fraction | Allelic Fraction | Genes mutated | Protein change | Nucleotide change | Allelic Fraction |
| 1 | IDH1 | NP_005887.2:p.Arg132Cys | NM_005896.3:c.394C>T | 0.4376 | 0.041 | NA | IDH1 | NP_005887.2:p.Arg132Cys | NM_005896.3:c.394C>T | 0.3706 |
| | NPM1 | NP_002511.1:p.Trp288CysfsTer12 | NM_002520.6:c.863_864insCAGA | 0.4319 | | | TP53 | NP_000537.3:p.Pro219Ser | NM_000546.5:c.655C>T | 0.0479 |
| | TP53 | NP_000537.3:p.Arg280dup | NM_000546.5:c.838_840dup | 0.1702 | | | | | | |
| | SF3B1 | NP_036565.2:p.Thr663Ile | NM_012433.3:c.1988C>T | 0.2899 | | | | | | |
| | CEBPA | NP_004355.2:p.Gly130Ser | NM_004364.4:c.388G>A | 0.1989 | | | | | | |
| | TET2 | NP_060098.3:p.Gln976ArgfsTer31 | NM_017628.4:c.2926del | 0.0105 | | | | | | |
| 44 | IDH2 | NP_002159.2:p.Arg140Gln | NM_002168.3:c.419G>A | 0.4313 | NA | 0 | | | | |
| | DNMT3A | NP_072046.2:p.Arg736His | NM_022552.4:c.2207G>A | 0.5565 | | | | | | |
| | ETV6 | NP_001978.1:p.Arg378Gln | NM_001987.4:c.1133G>A | 0.5153 | | | | | | |
| | ASXL1 | NP_056153.2:p.Gly646TrpfsTer12 | NM_015338.5:c.1934dup | 0.1212 | | | | | | |
| 45 | IDH1 | NP_005887.2:p.Arg132Cys | NM_005896.3:c.394C>T | 0.189 | 0 | 0 | | | | |
| | BCOR | NP_060215.4:p.Phe1566LeufsTer7 | NM_017745.5:c.4698_4699del | 0.2245 | | | | | | |
| | BCORL1 | NP_068765.3:p.Ser363PhefsTer55 | NM_021946.4:c.1088del | 0.1841 | | | | | | |
| | CUX1 | NP_001904.2:p.Ala422Gly | NM_001913.4:c.1265_1266delinsGG | 0.4506 | | | | | | |
| | DNMT3A | NP_072046.2:p.Arg882His | NM_022552.4:c.2645G>A | 0.1901 | | | | | | |
| | FLT3 | NP_004110.2:p.Leu576Pro | NM_004119.2:c.1727T>C | 0.1826 | | | | | | |
| 46 | IDH1 | NP_005887.2:p.Arg132His | NM_005896.3:c.395G>A | 0.0904 | 0 | 0 | | | | |
| | DNMT3A | NP_072046.2:p.Asn838Asp | NM_022552.4:c.2512A>G | 0.1155 | | | | | | |
| 47 | IDH1 | NP_005887.2:p.Arg132Cys | NM_005896.3:c.394C>T | 0.3444 | 0.0845 | 0 | | | | |
| | FLT3 | NP_004110.2:p.Asp593_Phe594insGlyPro | NM_004119.2:c.1779_1780insGGTCCC | 0.0956 | | | | | | |
| | DNMT3A | NP_072046.2:p.Ser775Pro | NM_022552.4:c.2323T>C | 0.3148 | | | | | | |
| | NPM1 | NP_002511.1:p.Trp288CysfsTer12 | NM_002520.6:c.860_863dup | 0.1268 | | | | | | |
| | TP53 | NP_000537.3:p.Arg273Leu | NM_000546.5:c.818G>T | 0.0162 | | | | | | |
| 48 | IDH1 | NP_005887.2:p.Arg132His | NM_005896.3:c.395G>A | 0.3592 | 0 | 0 | | | | |
| | PTPN11 | NP_002825.3:p.Gly503Ala | NM_002834.4:c.1508G>C | 0.2595 | | | | | | |
| | NPM1 | NP_002511.1:p.Trp288CysfsTer12 | NM_002520.6:c.860_863dup | 0.1341 | | | | | | |
| | FLT3 | NP_004110.2:p.Asp835Tyr | NM_004119.2:c.2503G>T | 0.0309 | | | | | | |

| | Baseline | | | | Pre-alloHCT Persistent IDH mutation | Post-alloHCT Persistent IDH mutation | Relapse | | | |
|---|---|---|---|---|---|---|---|---|---|---|
| Patient number | Genes mutated | Protein change | Nucleotide change | Allelic Fraction | Allelic Fraction | Allelic Fraction | Genes mutated | Protein change | Nucleotide change | Allelic Fraction |
| 1 | IDH1 | NP_005887.2:p.Arg132Cys | NM_005896.3:c.394C>T | 0.4376 | 0.041 | NA | IDH1 | NP_005887.2:p.Arg132Cys | NM_005896.3:c.394C>T | 0.3706 |
| | NPM1 | NP_002511.1:p.Trp288CysfsTer12 | NM_002520.6:c.863_864insCAGA | 0.4319 | | | TP53 | NP_000537.3:p.Pro219Ser | NM_000546.5:c.655C>T | 0.0479 |
| | TP53 | NP_000537.3:p.Arg280dup | NM_000546.5:c.838_840dup | 0.1702 | | | | | | |
| 49 | IDH2 | NP_002159.2:p.Arg140Gln | NM_002168.3:c.419G>A | 0.0706 | 0 | 0 | | | | |
| | RUNX1 | NP_001745.2:p.His404ProfsTer196 | NM_001754.4:c.1210dup | 0.0338 | | | | | | |
| | BCOR | NP_060215.4:p.Leu673PhefsTer67 | NM_017745.5:c.2018dup | 0.1405 | | | | | | |
| | STAG2 | | NM_006603.4:c.668-1G>A | 0.1437 | | | | | | |
| | EZH2 | NP_004447.2:p.Gln653Arg | NM_004456.4:c.1958A>G | 0.1523 | | | | | | |
| | SF3B1 | NP_036565.2:p.Lys700Glu | NM_012433.3:c.2098A>G | 0.1618 | | | | | | |
| 50 | IDH1 | NP_005887.2:p.Arg132His | NM_005896.3:c.395G>A | 0.1885 | 0 | 0 | | | | |
| | BCOR | NP_060215.4:p.Leu1552ProfsTer32 | NM_017745.5:c.4655del | 0.2339 | | | | | | |
| | WT1 | NP_000369.3:p.Ser364Ter | NM_000378.4:c.1091C>A | 0.2186 | | | | | | |
| 51 | IDH1 | NP_005887.2:p.Arg132Cys | NM_005896.3:c.394C>T | 0.3342 | 0 | 0 | | | | |
| | EZH2 | NP_004447.2:p.Pro486ArgfsTer26 | NM_004456.4:c.1457_1463del | 0.7623 | | | | | | |
| | CALR | NP_004334.1:p.Leu367ThrfsTer46 | NM_004343.3:c.1099_1150del | 0.4971 | | | | | | |
| | MYD88 | NP_002459.2:p.Asp288Asn | NM_002468.4:c.862G>A | 0.499 | | | | | | |
| | ASXL1 | NP_056153.2:p.Gly646TrpfsTer12 | NM_015338.5:c.1934dup | 0.3024 | | | | | | |
| 52 | IDH1 | NP_005887.2:p.Arg132His | NM_005896.3:c.395G>A | 0.3281 | 0.0089 | 0 | | | | |
| | PTPN11 | NP_002825.3:p.Asp61Ala | NM_002834.4:c.182A>C | 0.293 | | | | | | |
| | DNMT3A | NP_072046.2:p.Phe331Leu | NM_022552.4:c.991T>C | 0.3708 | | | | | | |
| 53 | IDH2 | NP_002159.2:p.Arg140Gln | NM_002168.3:c.419G>A | 0.3598 | 0.0437 | 0 | | | | |
| | SRSF2 | NP_003007.2:p.Pro95Arg | NM_003016.4:c.284C>G | 0.3717 | | | | | | |
| | FLT3 | NP_004110.2:p.Val491Leu | NM_004119.2:c.1471G>C | 0.0692 | | | | | | |
| | FLT3 | NP_004110.2:p.Asp835His | NM_004119.2:c.2503G>C | 0.1305 | | | | | | |
| | NPM1 | NP_002511.1:p.Trp288CysfsTer12 | NM_002520.6:c.860_863dup | 0.1498 | | | | | | |
| | FLT3 | NP_004110.2:p.Asp835Val | NM_004119.2:c.2504A>T | 0.0376 | | | | | | |
| 54 | IDH1 | NP_005887.2:p.Arg132His | NM_005896.3:c.395G>A | 0.1863 | 0.4596 | NA | | | | |

| | Baseline | | | | Pre-alloHCT Persistent IDH mutation | Post-alloHCT Persistent IDH mutation | Relapse | | | |
|---|---|---|---|---|---|---|---|---|---|---|
| Patient number | Genes mutated | Protein change | Nucleotide change | Allelic Fraction | Allelic Fraction | Allelic Fraction | Genes mutated | Protein change | Nucleotide change | Allelic Fraction |
| 1 | IDH1 | NP_005887.2:p.Arg132Cys | NM_005896.3:c.394C>T | 0.4376 | 0.041 | NA | IDH1 | NP_005887.2:p.Arg132Cys | NM_005896.3:c.394C>T | 0.3706 |
| | NPM1 | NP_002511.1:p.Trp288CysfsTer12 | NM_002520.6:c.863_864insCAGA | 0.4319 | | | TP53 | NP_000537.3:p.Pro219Ser | NM_000546.5:c.655C>T | 0.0479 |
| | TP53 | NP_000537.3:p.Arg280dup | NM_000546.5:c.838_840dup | 0.1702 | | | | | | |
| | IDH2 | NP_002159.2:p.Arg140Gln | NM_002168.3:c.419G>A | 0.2235 | 0.013 | NA | | | | |
| | SH2B3 | NP_005466.1:p.Ser337Cys | NM_005475.2:c.1010C>G | 0.3905 | | | | | | |
| | PHF6 | NP_115711.2:p.Ser248Pro | NM_032335.3:c.742T>C | 0.4403 | | | | | | |
| | PHF6 | NP_115711.2:p.Ser257Ter | NM_032335.3:c.770C>G | 0.0216 | | | | | | |
| | SMC1A | NP_006297.2:p.Arg807His | NM_006306.3:c.2420G>A | 0.3947 | | | | | | |
| | SRSF2 | NP_003007.2:p.Pro95His | NM_003016.4:c.284C>A | 0.4101 | | | | | | |
| 55 | IDH2 | NP_002159.2:p.Arg140Gln | NM_002168.3:c.419G>A | 0.1693 | NA | 0.0016 | | | | |
| | NPM1 | NP_002511.1:p.Trp288CysfsTer12 | NM_002520.6:c.860_861insTTGT | 0.0765 | | | | | | |
| | FLT3 | NP_004110.2:p.Pro606_Arg607insAsnGluTyrAspLeuLysTrpGluPhePro | NM_004119.2:c.1790_1819dup | 0.1085 | | | | | | |
| 56 | IDH2 | NP_002159.2:p.Arg172Lys | NM_002168.3:c.515G>A | 0.0041 | NA | 0 | | | | |
